# A Beta Regression Framework with Intentional Left-Censoring for Quantifying Familial Longevity

**DOI:** 10.64898/2026.01.13.26343996

**Authors:** Mar Rodriguez-Girondo, Niels van den Berg, Michel H. Hof

**Affiliations:** Department of Medical Statistics, Leiden University Medical Center, The Netherlands; Department of Molecular Epidemiology, Leiden University Medical Center, The Netherlands; Department of Epidemiology and Data Science, Amsterdam UMC, The Netherlands

## Abstract

Defining and quantifying exceptional familial human survival is a persistent challenge in longevity research. Traditional approaches rely on binary thresholds, arbitrary cutoffs, or simple descriptive measures, which discard information on variation among the oldest individuals, ignore differences in background mortality, and yield unstable family-level summaries. We propose a principled, model-based framework that transforms survival times into percentiles relative to population life tables, standardizing across birth cohorts, sexes, and populations. We extend beta mixed-effects regression to accommodate intentional left-censoring, which downweights early deaths while retaining their contribution to the likelihood, thereby focusing inference on extreme survival. Family-specific random effects provide interpretable, statistically grounded longevity scores, overcoming the limitations of ad hoc measures and enabling robust identification of long-lived families.

Simulation studies and application to a large multigenerational Dutch cohort demonstrate that the method reliably identifies families enriched for longevity. This framework provides a flexible, interpretable, and robust tool for analyzing familial survival, offering a paradigm shift in the statistical study of exceptional human lifespan.

## 1 Introduction

Understanding the causes of variation in human survival to extreme ages (longevity) is a central objective in aging research. Longevity and its determinants tend to cluster within families (van den Berg et al., 2017; Herskind et al., 1996; Perls et al., 2002; van den Berg et al., 2018a), making (multi-generational) family-based study designs essential for disentangling genetic and environmental influences (van den Berg, 2020a). Members of long-lived families often experience delayed onset of chronic and age-related diseases or even avoid them altogether (van den Berg et al., 2023; Andersen et al., 2012; Christensen et al., 2020). Studying families with exceptionally old individuals may therefore provide valuable insights into mechanisms that promote healthy aging and resilience to age-related conditions, ultimately informing strategies to extend healthspan in the general population (van den Berg, 2020a).

Identifying long-lived families presents both conceptual and methodological challenges. First, the definition of longevity itself is not trivial. Longevity is often treated as a binary trait, for example surviving to age 100 (Perls et al., 2002) or belonging to the top 10% of survivors within one’s birth cohort (van den Berg et al., 2020b; Rodriguez-Girondo et al., 2021). These definitions focus on survival to advanced ages, where the underlying protective mechanisms of aging are most likely to manifest. By excluding deaths at younger ages, often driven by external or disease-specific causes, binary definitions reduce mixing longevity-related mechanisms with those related to lifestyle and disease onset. However, an important limitation of binary definitions of longevity is that they impose arbitrary thresholds and discard valuable information on variation, especially above the cutoff, treating individuals who barely surpass the threshold as equivalent to those who live much longer. This loss of resolution can reduce statistical power, obscure meaningful biological differences, and complicate inference in the presence of right censoring, common in practice due to loss of follow-up. A more informative and flexible strategy is to conceptualize longevity as a continuous trait, down-weighting deaths at younger ages while retaining variation among the oldest individuals.

The second important challenge is that background mortality differs substantially between birth cohorts and countries (van den Berg et al., 2017). As a consequence, age is not an appropriate measurement scale when comparing survival across multiple generations and populations. Although it is possible to account for these differences through regression (Smith et al., 2009), standard models typically rely on strong assumptions that are often violated in this context. A more appropriate measure of survival is to use percentiles derived from publicly available life tables stratified by birth year, sex, and country. Using percentiles ensures comparability across cohorts; for example, two individuals born a century apart can be considered equally long-lived if both reach the 92nd percentile of their respective survival distributions, even if this corresponds to different chronological ages. With this transformation, the resulting survival percentile data are bounded between 0 and 1, and right-censoring due to loss to follow-up or survival beyond the observation window can be naturally represented by the last observed percentile combined with a censoring indicator.

A third challenge is that identifying long-lived families and summarizing familial longevity with a single score remains difficult. Simple descriptive measures, such as the proportion of long-lived relatives, are widely used but ignore differences in family size and estimation uncertainty (van den Berg, 2020a; Sebastiani et al., 2009). Importantly, this can lead to unstable family rankings. Robust family-level scores are needed for fair comparison between families and to provide reliable inputs for downstream (genetic) analyses.

To address these challenges, we propose an extension of the beta mixed-effects regression model for survival percentiles. Quantifying survival with percentiles naturally bounds the outcome between 0 and 1, making the beta distribution a flexible framework that can accommodate the skewness typical of survival at advanced ages. The beta mixed-effects model is extended to deal with non-informative left and right censoring. Left censoring is induced by a threshold. Under this scheme, individuals who die below the threshold are recorded as left-censored at the threshold percentile rather than as exact low-percentile observations. This effectively down-weights deaths at younger percentiles that are caused by factors unrelated to longevity, allowing us to focus on the extreme percentiles that reflect true longevity differences. The right censoring accounts for incomplete follow-up or survival beyond the observation window.

This model formulation gives family-specific random effects, which can be interpreted as longevity family scores. While mixed models have previously been applied for ranking purposes in hospital comparison studies (Lingsma et al., 2009), their use as a ranking tool in family studies, and in longevity research in particular, remains limited (Rodriguez-Girondo et al., 2021).

The rest of the paper is structured as follows: Section 2 presents the proposed methodology, including the likelihood formulation and the derivation of family longevity scores as empirical Bayes random effect estimates. Section 3 reports a simulation study designed to evaluate the ability of the proposed scoring approaches to detect long-lived families. Section 4 presents an illustration in the Historical Sample of the Netherlands (HSN), and Section 5 concludes with a discussion of the main findings, limitations, and potential extensions.

## 2 Beta regression for clustered survival percentile data

### 2.1 Survival percentile data

Deriving survival percentile data requires mapping individual survival ages to their corresponding percentiles in population-based life tables. This transformation standardizes survival relative to background mortality across birth cohorts, sexes, and populations, and yields outcomes bounded between 0 and 1.

Let *T*_0_ be the random survival time of interest given in age scale, *C* represent the right censoring process, assumed to be independent of *T*_0_. The observed time to death is given by the random variable *T* = min(*T*_0_, *C*) and Δ = *I*(*T*_0_ < *C*) is the non-censoring indicator. In our setting, we have access to life tables which provide population-based survival probabilities at age *t, t* ∈ [0, *t*_*max*_]. Denote the set of stratifying variables in the life table as **Z**. The set **Z** represents a collection of variables that are used to define distinct groups or strata within the population for which the life table is constructed. Typically life tables are birth-cohort- and sex-specific but other stratifying factors, like ethnicity and socio-economical status, are sometimes available.

Denote the available collection of life tables stratified by **Z** as *S*_0_(*t*, **Z**). This allows us to define a new random variable denoted by *Y*_0_ = *S*_0_(*T*_0_|**Z**), the survival percentile based on *T*_0_ given the life table *S*_0_(*t*, **Z**). Analogously, the censoring time *C* can be transformed into a censoring percentile *Y*_*C*_ = *S*_0_(*C*|**Z**). The observed survival percentile is defined as *Y* = min(*Y*_0_, *Y*_*C*_) and the non-censoring indicator Δ is also applicable in the percentile scale and can be rewritten as Δ = *I*(*Y*_0_ *< Y*_*C*_), indicating if *Y* refers to the actual survival percentile at death or at the last follow-up.

After determining the survival percentiles, we observe data from *n* families. For the *i*th family, consisting of *m*_*i*_ members, we observe

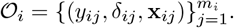

For the *j*th member of this family, *y*_*ij*_ = *S*_0_(*t*_*ij*_ | **z**_*ij*_) ∈ [0, 1] denotes the survival percentile and *δ*_*ij*_ a censoring indicator, where (*δ*_*ij*_ = 1 if the member died at *y*_*ij*_ and *δ*_*ij*_ = 0 if this member was still alive at *y*_*ij*_. In addition, **x**_*ij*_ = (*x*_1*ij*_, …, *x*_*qij*_) is a vector of *q* individual-specific covariates that may affect survival. Note that it is possible to include some of the stratifying variables used to obtain the life table *S*_0_(*t*, **z**_*ij*_) among these covariates.

### 2.2 Model for the percentiles

To model the percentiles, we use the regression parameterization of the beta distribution proposed by Ferrari and Cribari-Neto (Ferrari & Cribari-Neto, 2004). We add a latent family-specific random effect *U* that the unobserved familial longevity factors. We assume that *U* follows a normal distribution with mean zero and standard deviation *σ*, that is, *U* ∼*f*_*U*_ (*u*; *σ*) = *N* (0, *σ*^2^).

Conditional on *U* and covariates **X**, the survival percentile *Y* follows the beta distribution

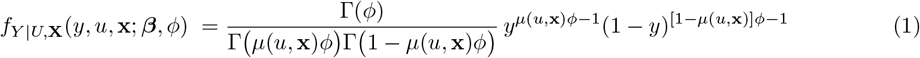

where the conditional mean is given by

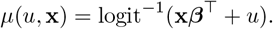

In addition, the normalization constant is given by

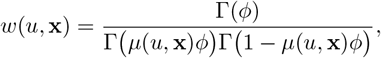

where Γ(*·*) is the gamma function.

The parameter vector ***β*** contains the associations between the covariates and the percentile outcome. In addition, the parameter *ϕ >* 0 is the precision parameter of the beta distribution. To illustrate the role of *ϕ*, note that the conditional variance of *Y* is

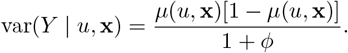

### 2.3 Censoring mechanisms and likelihood function

Before we estimate the parameters ***θ*** = (***β***, *σ, ϕ*) from the model describing the percentile outcomes, we have to address both left and right censoring of the outcome *Y*. Right censoring arises naturally in our sample because some individuals are still alive at the time of data collection or have missing death information. In contrast, left censoring is artificially imposed among the deceased individuals using a threshold *τ* ∈ (0, 1). Specifically, if *Y < τ* and *δ* = 1, we treat the percentile *Y* as left censored at *τ*.

Given these two censoring mechanisms, the observed data likelihood is

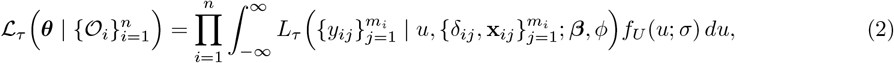

where the contribution of family *i*, conditionally on *U*, is given by

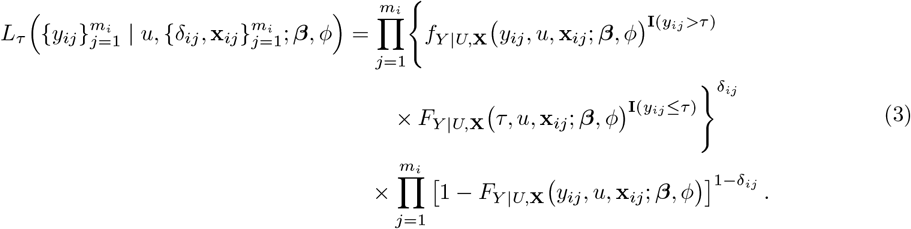

Here, **I**(·) denotes the indicator function, the conditional density function *f*_*Y* |*U*,**X**_(·) is given by Equation (1). The left-censored observations contribute to the likelihood via the conditional cumulative distribution function

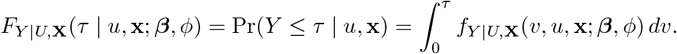

The threshold *τ* can be interpreted as a tunable focus parameter that controls how strongly inference is oriented to the upper tail of the percentiles. If this focus is not required, left censoring is omitted by setting *τ* = 0. In this case, survival percentiles are modeled continuously across the full range (0, 1). Importantly, all inference is conditional on the chosen threshold *τ*.

Rather than imposing left censoring at threshold *τ*, one might consider removing all individuals with survival percentiles below *τ*. However, removing those individuals can change the family-specific survival distribution and may artificially increase apparent familial longevity because members with shorter lifespans are ignored. Left censoring, by contrast, does not remove these individuals but down-weights their influence. As a result, the family structure is preserved, ensuring that inference targets biologically meaningful longevity signals.

An estimate of the parameter vector ***θ*** is obtained by maximizing the likelihood from Equation (2), i.e.

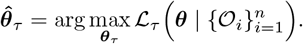

The integral with respect to the family-specific random effect *U* in the likelihood function is approximated numerically using adaptive Gaussian quadrature.

### 2.4 Family-Level Longevity Scores (FLBS and iFLBS)

The longevity family score for family *i* is defined as the expected family-specific random effect *U* conditional on the observed data and 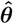. Using Bayes’ rule, this score is obtained as

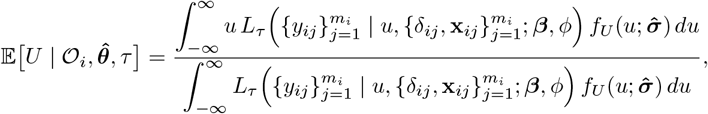

where the function *L*_*τ*_ (·) is given by Equation (3).

Our family-score offers a model-based summary of familial survival, accounting for family size, censoring, covariates, and random variability. Unlike descriptive summaries such as mean survival percentiles or counts of long-lived individuals, the family-score is corrected for these factors and stabilizes inference across unequal family sizes and incomplete follow-up, enabling coherent comparisons across families.

We define two versions of the family-score: the Family-Level Beta Scores (FLBS) and the informed Family-Level Beta Scores (iFLBS). For family *i*, both scores are defined as

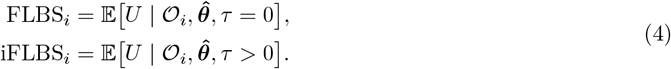

The iFLBS inherits the intentional left-censoring structure introduced in Section 2.3, thereby directing inference toward the upper tail of the survival distribution. By downweighting early deaths, the iFLBS captures longevity-specific familial tendencies while preserving the full information in the likelihood. Together, both scores enable a coherent comparison of familial longevity patterns under varying degrees of longevity emphasis, with the threshold *τ* serving as a flexible parameter for tuning the biological focus of inference.

## 3 Simulation study

### 3.1 Simulation setup

#### 3.1.1 Aim and estimands

To evaluate the new beta mixed-effects model approach for multi-generational survival data in longevity research, we performed a simulation study focused on identifying long-lived families. Our primary interest was the family-specific latent component, which captured the degree of familial aggregation of longevity. In particular, the objective was to accurately identify the 10% of families with the strongest underlying predisposition to longevity.

#### 3.1.2 Data generation

For each simulated family *i*, the number of members was sampled from a discrete uniform distribution on the values {2, 3, …, 8}. Familial aggregation of longevity was represented by a family-specific random effect *U*_*i*_ ∼ *N* (0, *σ*^2^), where *σ* = 0.5.

For each individual *j* in family *i*, the survival percentile *y*_*ij*_ was drawn from the mixture distribution

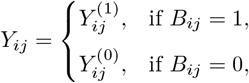

where individuals either experience survival outcomes caused by familial longevity or to external, non-familial factors. Specifically, 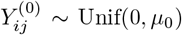 represents variation due to external factors. In addition, 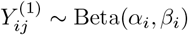 describes the familial longevity, where the family-specific parameters are given by *α*_*i*_ = *µ*_*i*_*ϕ* and *β*_*i*_ = (1 − *µ*_*i*_)*ϕ*. Here, the family-specific mean is given by

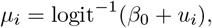

where *β*_0_ denotes a fixed effect and *u*_*i*_ is the family-specific random effect. The probability that the survival percentile is caused by familial longevity is given by Pr(*B*_*ij*_ = 1) = *π*.

We considered two scenarios with respect to *π*. In Scenario 1, we set *π* = 1, meaning that all survival percentiles were driven by the familial longevity component. In Scenario 2, we set *π* = 0.5, such that half of the individuals in each family die from non-familial causes. With respect to the parameters, we set *µ*_0_ = 0.9, *β*_0_ = 2.2 and *ϕ* = 20 in both scenarios. These parameters lead to a realistic distribution of survival percentiles with mean 0.7, median 0.8 and 0.9 and the 75th percentile at 0.9 in Scenario 2. The choice of *µ*_0_ = 0.9 reflects evidence from the longevity literature indicating that external causes contribute little to survival at higher percentiles (Evans et al., 2014; Sebastiani et al., 2015). For each scenario and Monte Carlo trial, we simulated data for *n* ∈ {200, 400} families and we assumed 30% of observations subject to independent uniform right-censoring on the interval [0, 1].

#### 3.1.3 Methods and performance measures

We calculated both the FLBS and the iFLBS scores, given by Definition (4). For the iFBLS, the threshold was set at the 90th percentile, that is, *τ* = 0.9. The FLBS summarizes family-level survival percentiles across the entire distribution, accounting for family size, random effects, and censoring, whereas the iFLBS emphasizes the upper tail of the survival distribution by downweighting early deaths below the threshold *τ*.

To assess the utility of the methods for selecting long-lived families, we repeated the simulation *M* = 500 times and computed the positive predictive value (PPV) in each Monte Carlo trial. Families with scores in the top 10% of the score distribution were classified as long-lived. The PPV was defined as the proportion of these selected families whose underlying simulated random effect *u* also fell within the top 10%, providing a direct measure of the accuracy of the FLBS and iFLBS in detecting genuinely long-lived families.

### 3.2 Simulation results

The main results are presented in Figure 1. In Scenario 1, FLBS and iFBLS exhibit similar performance, with mean PPV values of approximately 0.5 for both sample sizes considered. The Monte Carlo standard errors decrease with increasing sample size, from 0.09 at *n* = 200 to 0.06 at *n* = 400, for both methods. In contrast, in Scenario 2, iFBLS clearly outperforms the FLBS. The mean PPV across the *M* = 500 Monte Carlo trials is 0.2 for FLBS at both sample sizes, with Monte Carlo standard errors of 0.10 in both cases. The mean PPV for iFBLS is 0.4, with Monte Carlo standard errors of 0.10 and 0.06 for the two sample sizes *n* = 200 and *n* = 400, respectively. These results support the effectiveness of the informed method in the more realistic setting where unrelated deaths occur at younger ages. Interestingly, the thresholded approach remains robust when the underlying data-generating mechanism is fully driven by the familial random effect, which determines age at death across the entire lifespan. The intentional left-censoring strategy proves particularly valuable, as it allows the method to focus on complex longevity mechanisms at the oldest ages while still performing well in more agnostic settings where high-age emphasis is less clear.

**Figure 1.**
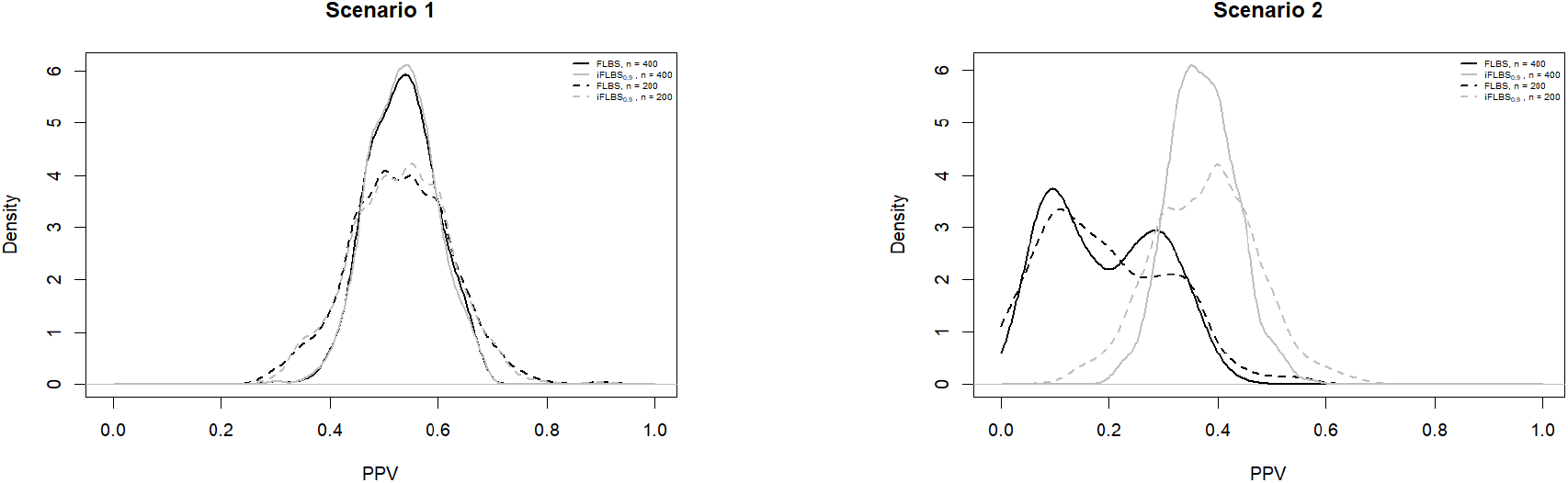
Simulation Study. Positive predictive values (PPV) of FLBS and iFLBS with *τ* = 0.9 (denoted by iFLBS_0.9_) computed over *M* = 500 Monte Carlo trials. Families with scores in the top 10% of the score distribution were classified as long-lived. The ositive predictive value (PPV) is defined as the proportion of these selected families whose underlying simulated random effect *u* also lies in the top 10% of its true distribution. Left panel: Scenario 1. Right panel: Scenario 2.

## 4 Real data application

We applied the proposed longevity scores on the data from the Historical Sample of the Netherlands (HSN), a multigenerational family dataset of Dutch individuals born between 1850 and 1922 (Mandemakers, 2000; van den Berg et al., 2018b). The dataset contains lifetime information on the members of 1326 five-generational families, each centered around a single proband (Mandemakers & Munnik, 2016; van den Berg et al., 2020b). For this analysis, we constructed familial longevity scores from descendants in the second (F2) and third (F3) generations. The resulting subsample included 1241 families with 19,928 participants born between years 1879 and 1973. The follow-up ended in 2017, resulting in a right-censoring rate of 30%.

We focused primarily on the FBLS and the iFBLS with varying cut-offs at the 70%, 80% and 90% percentiles (*τ* = 0.7, 0.8, 0.9). We also considered a alternative approach based on a Cox proportional hazards model with shared frailty, adjusted for sex and year of birth. The resulting frailty estimates from this model were used as family-level scores. Note that we only focused on methods that can appropriately handle right-censored data, which excludes methods based on binary definitions of longevity.

To evaluate the utility of the scores as selection tools for longevity studies, we performed an internal validation. Although a full validation would require comprehensive genetic and environmental data to fully characterize what these scores reflect, this internal validation provides an initial demonstration of their practical value in multi-generational survival data. The internal validation consisted of randomly selecting one individual per family and excluding them from the score calculation. We then simulated selections based on family longevity, defining long-lived families as those with a family score in the top 10% of the distribution. Finally, we applied Cox proportional hazards regression, adjusting for sex and year of birth, to assess the survival advantage of individuals from these high-scoring families.

The results from the internal validation have been summarized in Figure 2. The thresholded iFLBS at the 70th and 80th percentiles (*τ* = 0.7, 0.8) were strongly associated with lower mortality risk, yielding nearly identical estimates (HR = 0.75, 95% CI: 0.60–0.93). The 90th percentile threshold (*τ* = 0.9) also showed a protective effect (HR = 0.80, 95% CI: 0.64–0.99), though slightly weaker. In contrast, the FLBS and the Cox-based frailty score did not reach statistical significance. These results support the use of thresholded iFLBS for identifying long-lived families in longevity research.

**Figure 2.**
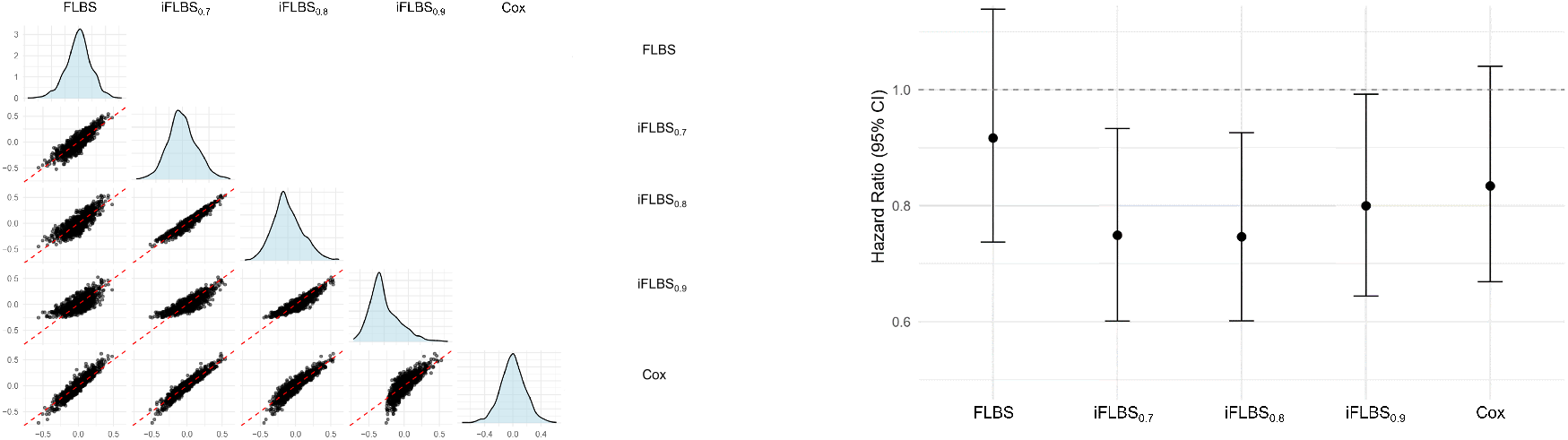
Real data application. Left panel: Distribution and pairwise visual comparison (via scatterplots) of the five considered scores: FLBS, iFLBS at the 70th and 80th and 90th percentiles, denoted by iFLBS_0.7_, iFLBS_0.8_, and iFLBS_0.9_ respectively, and the frailty Cox-based score, denoted by Cox. Right panel: Hazard ratios (HRs) and 95% confidence intervals for the association between family-based longevity scores (top 10% selection) and individual survival, adjusted for sex and year of birth. The dashed line at HR = 1 indicates no survival advantage.

Defining long-lived families as those with scores in the top 10% for each method results in only modest overlap: 53 out of the 1,241 families were identified as long-lived by all five scoring methods. This indicates that more than half of the top selections are method-specific, despite substantial overall correlation across the scores (see the left panel of Figure 2).

## 5 Discussion

In the longevity literature, extreme survival is often regarded as a distinct biological trait that could potentially reveal protective survival and health mechanisms that differ from those determining general lifespan. A central challenge is that chronological age is a population-dependent measure: an age that is ordinary in one context may represent exceptional survival in another. For example, in high-mortality populations, a man who survives to age 85 may belong to the top few percent of his cohort’s survival distribution, biologically comparable to reaching age 100 in a low-mortality population. Analyses based on fixed age thresholds cannot account for these differences, as the same absolute age reflects vastly different survival rarity across populations.

To address this limitation, our framework models survival percentiles rather than absolute ages. Percentiles express survival relative to a cohort’s mortality distribution, automatically standardizing for secular trends, environmental conditions, and sex- or region-specific hazards. Familial clustering can then be interpreted as resilience relative to peers, independent of background mortality.

Building on this idea, we introduced a beta regression model with intentional left-censoring, a mechanism that operationalizes the conceptual distinction between lifespan and longevity. The left-censoring threshold downweights variation among short-lived individuals, treating them as equally “non-long-lived,” and concentrates inference on the upper tail of the survival distribution, where longevity-related genetic or environmental signals are most informative. This targeted left-censoring functions as a penalization term within a coherent likelihood framework, allowing the analyst to tune the model’s focus on extreme survival while retaining full probabilistic interpretability. It should be noted that simply removing low-survival individuals is not appropriate. These individuals do not contribute information about extreme longevity, but excluding them would alter the family’s survival profile and could make the family appear more long-lived than it actually is. By contrast, left-censoring retains all individuals in the likelihood, while downweighting their influence on the upper tail, ensuring that inference focuses appropriately on biologically meaningful longevity signals.

Simulation studies confirmed that the informed, left-censored family-level beta score yield stable and biologically meaningful rankings across varying sample sizes and right-censoring levels. It effectively identifies truly long-lived families under scenarios where clustering occurs only at extreme survival percentiles. Application to a large multigenerational Dutch cohort further validated the approach: informed left-censored family-level beta scores highlighted families with clear survival advantages that were less apparent under an uninformed model.

Although there is general consensus that longevity should be regarded as a distinct trait, the use of percentiles versus absolute extreme age remains subject to debate. An alternative view emphasizes the use of very high ages to define exceptional survival (Andersen et al., 2012), implicitly assuming that longevity mechanisms can only be identified at the highest attainable ages. Which perspective is more appropriate is arguable, but in both cases, the construction of meaningful family-level scores necessarily requires a principled way of downweighting early deaths and emphasizing late survival. Our intentional left-censoring approach is compatible with both views, as it provides a systematic framework for tuning the degree of emphasis on extreme survival to be tuned beyond ad hoc binary cut-offs and could also be applied within hazard models based on the age scale.

Beyond longevity research, our proposal is directly applicable to other multigenerational settings where risk is defined relative to a population distribution rather than on an absolute age scale. A prominent example is hereditary cancer (Brandt et al., 2008; Litton et al., 2012), where ages at onset are subject to right censoring and vary by sex, birth cohort, screening practices, and background incidence. In such settings, absolute age thresholds are difficult to interpret across families and generations, whereas percentiles provide a standardized measure of relative extremeness. Here, the uninformed version of our model without left-censoring offers a natural tool for quantifying familial clustering of unusually early onset while properly accounting for right censoring.

Future extensions could include smoother weighting functions instead of discrete left-censoring to provide additional flexibility. Although our analyses focused on unadjusted family-level scores, the model can readily accommodate covariates; future applications could adjust for environmental factors such as socioeconomic status to isolate the genetic component of longevity. This decomposition is not feasible with simple descriptive metrics, underscoring a key advantage of a model-based approach.

In conclusion, our framework brings together several methodological innovations to provide a principled and coherent approach to studying familial longevity. By expressing survival on the percentile scale, we achieve comparability across birth cohorts, sexes, and populations while retaining the full information of continuous survival data. Extending beta mixed-effects regression to incorporate both right and intentional left censoring addresses long-standing challenges in differentiating early deaths from genuinely exceptional survival, offering a rigorous solution that has been lacking until now. The resulting model-based family scores provide a statistically grounded and interpretable summary of familial longevity, overcoming limitations of ad hoc descriptive measures. Taken together, these contributions enable more robust identification of families enriched for exceptional survival and support biologically meaningful inference in multi-generational studies.

## Data Availability

The HSN data are publicly available at https://hdl.handle.net/10622/MVUAIP

## Notes

### Competing Interest Statement

The authors have declared no competing interest.

### Funding Statement

Netherlands Organization for Scientific Research, domain Health Research and Medical Sciences (09120012010052)

### Author Declarations

The HSN data are publicly available at https://hdl.handle.net/10622/MVUAIP

